# Artificial intelligence-based ECG reconstruction error as a continuous predictor of all-cause mortality: a multi-cohort retrospective validation study

**DOI:** 10.64898/2026.07.10.26357749

**Authors:** Angus Nicolson, Samuel Pröll, Riccardo Lunelli, Hagen Blankenburg, Peter Pramstaller, Christian Fuchsberger, Axel Bauer, Clemens Dlaska

## Abstract

**Background:** Recent artificial intelligence (AI) models applied to the electrocardiogram (ECG) for risk stratification typically rely on supervised learning, defining risk as the error relative to an external target such as age or sex. This couples the risk score to the choice of target rather than the cardiac signal alone, and may limit generalisability. We aimed to develop a self-supervised AI-ECG risk score based on the error in reconstructing a partially masked ECG.

**Methods:** A transformer-based masked autoencoder was trained on 85% of the CODE dataset (n = 7,212,109 ECGs) to reconstruct ECG signals from partially masked inputs. The association between reconstruction error and all-cause mortality was assessed internally in CODE-15% and externally validated in four independent cohorts: MIMIC-IV-ECG (critical care, US), HEEDB (hospital, US), CHRIS (population-based, Italy), and Innsbruck (cardiology centre, Austria). A binary risk score (>1 SD above the CODE-15% mean) was additionally evaluated in these cohorts and in the UK Biobank (population-based, UK).

**Findings:** In Cox proportional hazards models adjusted for age and sex, each 1-SD increase in reconstruction error was associated with higher all-cause mortality (all p<0.001; cohort median follow-up 1.4-11.0 years): CODE-15% (HR 1.39, 95% CI 1.37-1.42), MIMIC-IV-ECG (HR 1.39, 95% CI 1.37-1.40), HEEDB (HR 1.41, 95% CI 1.40-1.41), Innsbruck (HR 1.23, 95% CI 1.21-1.26), and CHRIS (HR 1.25, 95% CI 1.14-1.38). The binary threshold identified a high-risk group with increased mortality in all six cohorts, including the UK Biobank (HR 1.27, 95% CI 1.08-1.50, p=0.004).

**Interpretation:** Reconstruction error is a generalisable predictor of all-cause mortality across diverse clinical and population-based settings. Unlike supervised approaches, it reflects the model’s uncertainty about the ECG signal itself rather than error relative to an external target, providing a direct measure of how much each recording deviates from normal cardiac electrical patterns.

## Introduction

Cardiovascular disease (CVD) remains the leading cause of mortality worldwide, accounting for over 19.2 million deaths annually^1^. Early identification of individuals at elevated cardiovascular risk is central to prevention strategies, yet existing risk scores depend on clinical variables that may be unavailable, inaccurately measured, or poorly predictive at the individual level.^2–4^ The 12-lead ECG remains one of the most widely used diagnostic tools in cardiovascular medicine, owing to its low cost, non-invasiveness, and near-universal availability.

Recent advances in deep learning have enabled neural networks to extract prognostically relevant information directly from raw ECG waveforms, surpassing conventional ECG interpretation based on predefined intervals and amplitudes^5–7^. Supervised AI-ECG models have been shown to predict age and sex^8^, incident atrial fibrillation^9^, and heart failure^10^. Notably, the discrepancy between chronological age and AI-predicted ECG age (delta age) has been shown to be independently predictive of mortality^11–15^. However, supervised approaches of this kind may learn cohort-specific or outcome-specific artefacts, potentially limiting generalisability.

Self-supervised learning offers an alternative paradigm by learning representations from the intrinsic structure of the data rather than from externally defined labels. Masked autoencoders (MAEs), originally developed for the computer vision and natural language domains, learn by reconstructing missing portions of the input signal^16,17^. Reconstruction error (the discrepancy between the original and reconstructed signal) has been established as an effective measure for anomaly detection across diverse domains including industrial applications^18^, medical imaging, and ECG signals^19,20^. In this work, instead of anomaly detection, we propose using reconstruction error as a direct measure of all-cause mortality risk.

When applied to ECGs, the masking process of an MAE has a specific and interpretable meaning. Each of the 12 ECG leads represents a distinct spatial projection of the heart’s three-dimensional electrical dipole onto a particular anatomical axis. In our implementation, reconstruction error is computed with 80% of the ECG patches masked, under the constraint that no timepoint is entirely unobserved: the model always has access to at least one lead at any given timepoint. The reconstruction task therefore requires the model to predict, for each timepoint, what the remaining leads would show. A model trained on a large population of ECGs will learn the characteristic temporal and cross-lead patterns that define normal cardiac electrical activity. Reconstruction error reflects the degree to which an ECG deviates from these learned patterns. Critically, this signal is entirely intrinsic to the ECG itself. The error does not depend on any participant demographic information, clinical label, or external reference value. This distinguishes it from supervised approaches such as ECG-age, where risk is defined as the discrepancy between a model prediction and an externally provided ground truth. The reconstruction error is a direct, label-free measure of the model’s uncertainty about the cardiac electrical field.

We assessed the prognostic value of this reconstruction-based risk score for all-cause mortality across six cohorts spanning primary care, critical care, hospital, cardiology, and population-level settings, and evaluated its generalisability independent of clinical covariates and standard ECG parameters.

## Methods

### Study Population and Data Sources

This is a retrospective, observational, multi-cohort validation study. The model was trained using ECGs from the CODE dataset^7,21^, derived from the Telehealth Network of Minas Gerais (TNMG), a public telehealth system serving 811 of 853 municipalities across Minas Gerais, Brazil. ECGs were recorded predominantly in primary care facilities between 2010 and 2016, with recording durations of 7-10 seconds and sampling frequencies of 300–600 Hz. The full CODE dataset comprises eight million ECGs from two million patients^7,21^. External validation was performed in cohorts selected to represent diverse healthcare settings and populations (Figure 1d and Figure 1e).

**Figure 1:**
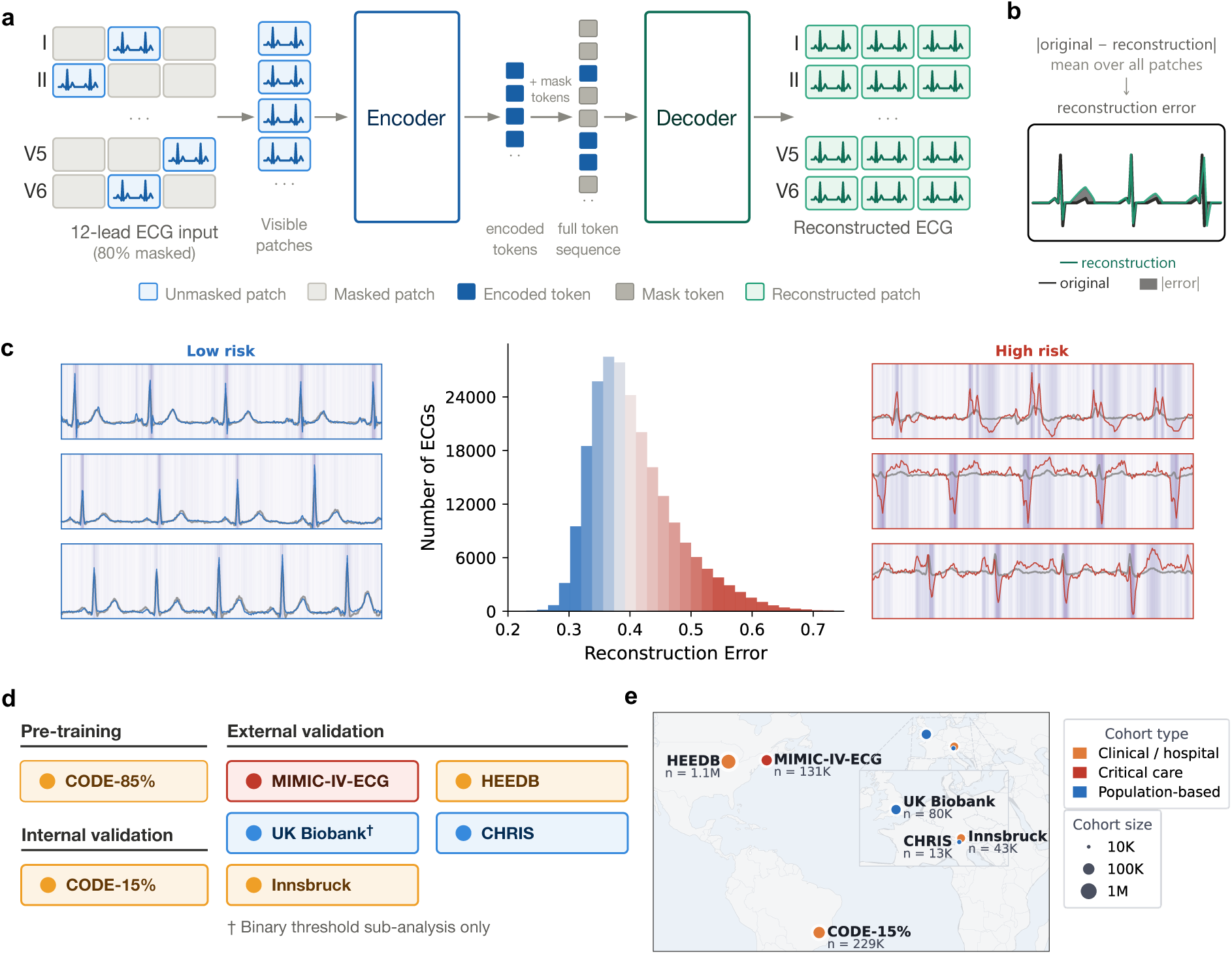
Reconstruction error based ECG risk score. **a** The ECG is split into 36 patches, 80% of the patches are masked and the remaining patches are passed to the ViT encoder. After the mask tokens are added (indicating which regions were removed), the decoder reconstructs the original ECG. **b** Reconstruction error is calculated as the mean absolute error between the reconstruction and the original ECG. **c** higher reconstruction error is associated with abnormal ECG patterns and increased risk of all-cause mortality **d** The model was pretrained on 85% of CODE. Reconstruction error was internally validated as a predictor of all-cause mortality on CODE-15% and externally validated on MIMIC-IV-ECG, HEEDB, CHRIS, Innsbruck, and the UK Biobank (binary sub-analysis only). **e** The validation cohorts span three continents and 1.6M participants.

CODE-15%,^11^ comprising a single ECG from 15% of patients from the full CODE dataset stratified by age, was withheld from model training (n = 229,499 ECGs). CODE-15% served as the primary internal validation cohort, the dataset used to normalise the reconstruction errors of the other cohorts, and was used to define the binary risk threshold used in sub-analyses. All ECGs from patients in CODE-15% were excluded from model training. External validation cohorts used in the primary analysis comprised of:

1. MIMIC-IV-ECG,^22,23^ comprising ECGs from critically ill and emergency department patients at the Beth Israel Deaconess Medical Center, Boston, USA, and characterised by a high burden of acute and chronic illness and elevated short-term mortality.
2. HEEDB (Harvard-Emory ECG Database),^24^ a large US hospital-based ECG dataset from multiple centres, representing a broad clinical spectrum of inpatient and outpatient presentations.
3. CHRIS (Cooperative Health Research in South Tyrol),^25,26^ a population-based cohort recruited from 13 municipalities in the alpine Val Venosta/Vinschgau district in the Bolzano/Bozen - South Tyrol province of northern Italy.
4. Innsbruck,^15^ a cardiology cohort from the Medical University of Innsbruck, Austria, comprising inpatient, outpatient, and emergency department patients with a high prevalence of acute and chronic cardiovascular disease.

Inclusion criteria were age ≥18 years and availability of at least one standard 12-lead ECG greater than 7s in duration. During validation, for individuals with multiple ECGs, only the first recording was used to avoid informative censoring. During model training, all ECGs for each participant were used. For the primary analyses, ECGs were included regardless of rhythm (sinus rhythm, atrial fibrillation, pacemaker) or level of noise. Ethical approval for each dataset was obtained by the respective institutions. All analyses were conducted using pre-existing, de-identified data.

The UK Biobank,^27^ a prospective population-based cohort of volunteers (aged 40–69 years at enrolment) recruited from across the United Kingdom, was additionally used to validate a binary threshold of reconstruction error. Due to current restrictions on data access^28^, the UK-Biobank results are incomplete and only the hazard ratio for this sub-analysis is available.

### Endpoints and Follow-up

The primary endpoint was all-cause mortality. Follow-up time was defined from the date of ECG acquisition to the occurrence of the endpoint, censoring, or end of follow-up. For most cohorts, mortality information was obtained from national death registries, hospital records, or linkage to electronic health records. For CHRIS, mortality information was obtained from local newspaper and church obituaries and by direct contact with participants’ families. Participants in CHRIS may withdraw consent for data use after death, in which case their records are destroyed. For primary analyses the maximum available follow-up time for each cohort was used. Both the UK Biobank and CHRIS have continuous follow-up, with mortality information updated regularly. The mortality data for the UK Biobank was last updated 31^st^ August 2024 for participants from England and Wales and 30^th^ November 2024 for participants from Scotland. The mortality information for CHRIS was last updated on 24^th^ June 2026. Median follow-up durations and event rates are summarised in Table 1.

**Table 1:**
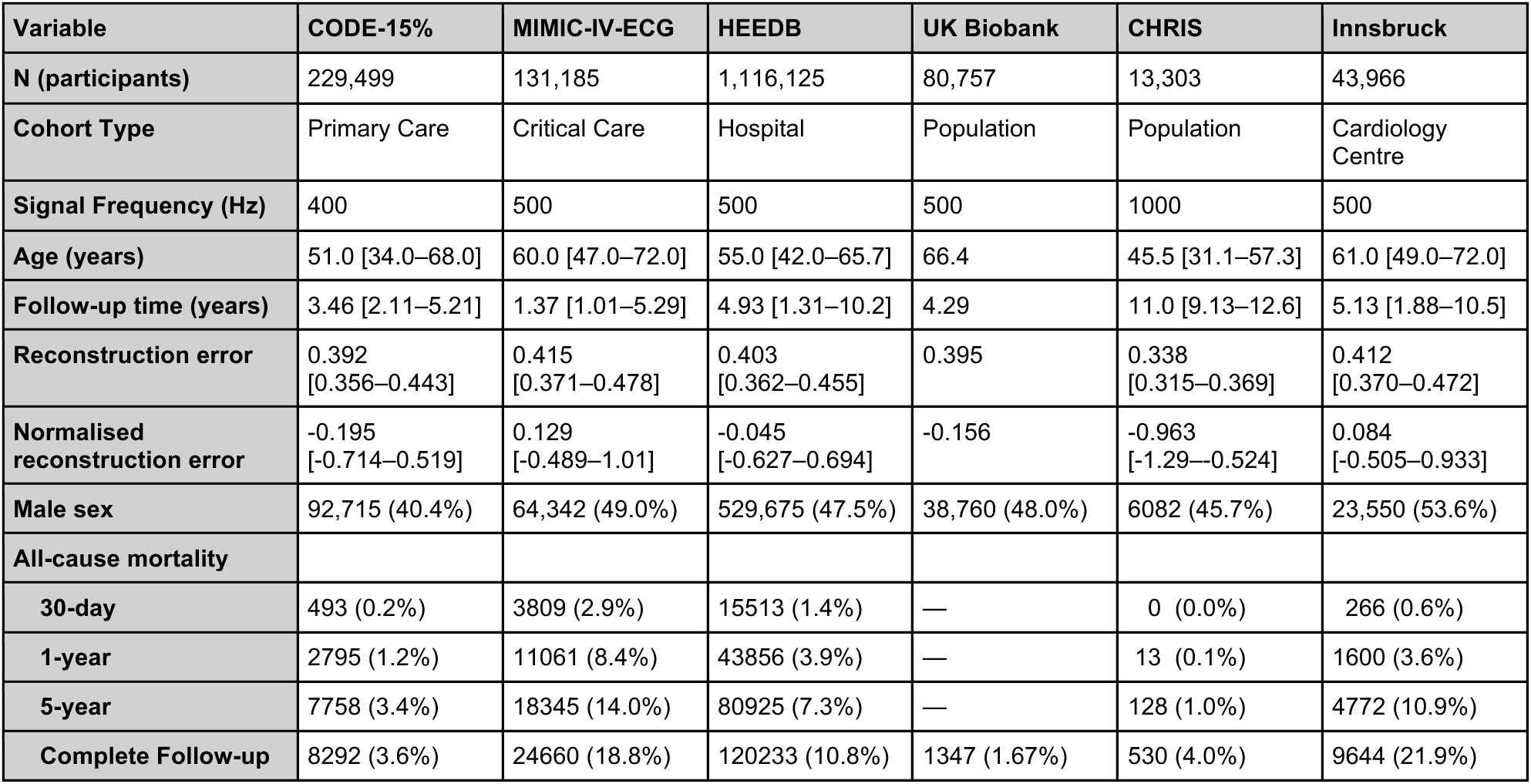
Demographic information for each cohort. All values are for the cohorts after inclusion/exclusion criteria are applied. For continuous variables the median and inter-quartile range (IQR) are displayed. For binary variables, the number of participants and proportion of the cohort are shown. Due to current restrictions on data access^28^, the UK-Biobank metrics are incomplete.

### AI Model Development

ECGs were resampled to 100 Hz, and each lead was normalised separately to have a mean of zero and a standard deviation of one. This normalisation removes information about differences in absolute amplitude between leads, encouraging the model to focus on waveform morphology rather than absolute voltage. The signals were filtered using a 4^th^ order bandpass (0.5-40 Hz) Butterworth filter. Stronger filtering and resampling to a lower frequency were chosen to ensure similar ECG characteristics across acquisition sites, enabling the risk score threshold derived from CODE-15% to generalise to other datasets acquired under different conditions.

The model architecture consisted of a vision transformer (ViT) adapted for ECG signals (see Figure 1a for an overview).^29,30^ For each forward pass of the model, a random segment of 7.68 s of the ECG was cropped. This length was chosen because CODE recordings range from 7–10 seconds, minimising the need for padding, and because 7.68 seconds is exactly divisible into patches of 256 samples at 100 Hz. Each lead was split into three non-overlapping patches of length 256 samples (2.56 seconds), giving 36 patches in total across a 12-lead ECG. During training, 80% of the patches were randomly masked, and the model was trained to reconstruct the original waveform from the visible segments using a mean squared error loss applied only to the masked regions. The loss function was not applied to regions exactly equal to zero (padded signals). The model architecture was composed of a ViT-Tiny encoder (12 layers) and a shallower decoder (4 layers). During training we used an AdamW optimiser^31^, a learning rate of 0.0005 with 1 epoch of warmup and a cosine annealing schedule, and a batch size of 2048 for a maximum of 100 epochs. The model was pre-trained on 85% of the CODE dataset (n = 7,212,109 ECGs).

To identify the optimal model for risk stratification, we systematically evaluated models of varying capacity (ViT-Tiny, ViT-Small, and ViT-Base) and at multiple checkpoints throughout training, assessing both reconstruction quality and prognostic performance. This evaluation was initially performed on the validation set only (CODE-15%). This analysis found that larger models reconstructed ECG signals with greater fidelity but with inferior risk stratification performance. Hence, we use the smallest model (ViT-Tiny) as the primary model, which also offers practical advantages in terms of inference speed. The main results reported in this paper are from a ViT-Tiny model that was trained for only 2600 steps (slightly less than 1 epoch of training). Once the primary model was chosen, these analyses were repeated for MIMIC-IV-ECG, Innsbruck and HEEDB, to demonstrate that the patterns generalise across cohorts.

### Risk Score Derivation

We quantified reconstruction error using the mean absolute error between the original and reconstructed ECG across all leads and time points (Figure 1b). Each ECG is processed 5 times with a different random 7.68 s crop and a different set of masked patches. Unlike during training, the masking at inference is constrained so that no timepoint was left completely masked. This ensures the model always has knowledge of the location of the R-peaks and reduces variance in the repeat measurement of a signal. The mean error across the five repeated inference runs is referred to as the reconstruction error throughout the paper.

We use the mean (0.406) and standard deviation (0.071) of the CODE-15% reconstruction error to z-score normalise the reconstruction error of each cohort. This expresses each value in standard deviations from the CODE-15% mean, giving an interpretable and consistent scale for Cox proportional hazards modelling across cohorts.

As a sub analysis, we defined a binary risk score by applying a threshold of 1.0 to the normalised reconstruction error. Participants with a normalised error above 1.0, equivalent to a raw error above the CODE-15% mean plus one standard deviation (0.477), were classified as high risk and all others as low risk. The sensitivity of the method to this threshold was assessed and is reported in the supplementary.

### Statistical Analysis

All analyses were performed in Python. Associations between the risk score and outcomes were assessed using Cox proportional hazards models. Unless otherwise stated, models were fit adjusted for age and sex and for the maximum available follow-up time for each cohort (no additional right censoring). Hazard ratios for reconstruction error are for the normalised reconstruction error, meaning the hazard is for an increase of one standard deviation of CODE-15% (0.071). Reconstruction error was entered as a continuous covariate in all primary models.

In cohorts with at least one death within the specified time frame, additional Cox models were fit for 30-day, 1-year, and 5-year mortality. On MIMIC-IV-ECG an additional model was fit adjusting for standard ECG machine-read measures and on Innsbruck an additional model was fit adjusted for comorbidities. Discrimination was assessed using the Harrell’s C-index and time-dependent receiver operating characteristic (ROC) curves, calculated at 30 days, 1 year, and 5 years. Kaplan-Meier survival curves were used to visualise differences in survival between participants from the different quartiles of reconstruction error within each cohort. Statistical significance of hazard ratios for individual covariates was ascertained using the Wald test. Statistical significance was defined as p<0.05.

Sub-group analyses were performed in MIMIC-IV-ECG to assess robustness of the risk score across clinically defined patient subsets: patients with good-quality ECGs (as defined by automated signal quality assessment), patients without pacemakers, patients with valid machine-read ECG parameters, patients without recorded premature ventricular contractions (PVCs), and patients without conduction defects. Conduction defect labels were defined using mentions of the following in the machine-generated reports: bundle branch block, fascicular block, intraventricular conduction defect (IVCD), or WPW syndrome.

### Model Explainability

All model explainability results were performed on a random sample of 5000 patients from MIMIC-IV-ECG. We sample ECGs from the top and bottom decile of reconstruction error to provide a qualitative overview of the types of ECGs that the model deems low/high risk. For quantitative comparisons between risk groups, we instead used the binary risk threshold defined above. We report the mean error across each lead for the high- and low-risk groups, highlighting differences in the distribution of error between them. Additional analyses were performed on individual ECG segments/intervals. We used a previously trained ECG segmentation model (a U-Net^32^ trained on PTB-XL+^33^) to segment the P wave, T wave and QRS complex for each ECG. Using these three segmentations we define the following regions: P wave, PQ segment, QRS complex, ST segment, T wave, TP interval, and summed the contribution of each region to the total reconstruction error.

## Results

Baseline characteristics of all cohorts are summarised in Table 1. The datasets covered a wide spectrum of participants and study characteristics, with the median participant age ranging from 45.5 years (CHRIS) to 66 years (UK Biobank), the median follow-up period ranging from 1.37 years (MIMIC-IV-ECG) to 11.0 years (CHRIS), and the 5-year mortality rate ranging from 1.0% (CHRIS) to 14.0% (MIMIC-IV-ECG).

The primary results are displayed in Figure 2. In Cox proportional hazard models adjusted for age and sex, an increase in reconstruction error was significantly associated with increased all-cause mortality across all cohorts (Figure 2b). Hazard ratios (HRs) were greatest in clinical and hospital-based cohorts: MIMIC-IV-ECG (HR 1.39, 95% CI 1.37-1.40, p<0.001, C-index 0.731), HEEDB (HR 1.41, 95% CI 1.40-1.41, p<0.001, C-index 0.740), and CODE-15% (HR 1.39, 95% CI 1.37-1.42, p<0.001, C-index 0.810). Although attenuated, a significant association was also observed in Innsbruck (HR 1.23, 95% CI 1.21-1.26, p<0.001, C-index 0.741) and CHRIS (HR 1.25, 95% CI 1.14-1.38, p<0.001, C-index 0.896).

**Figure 2:**
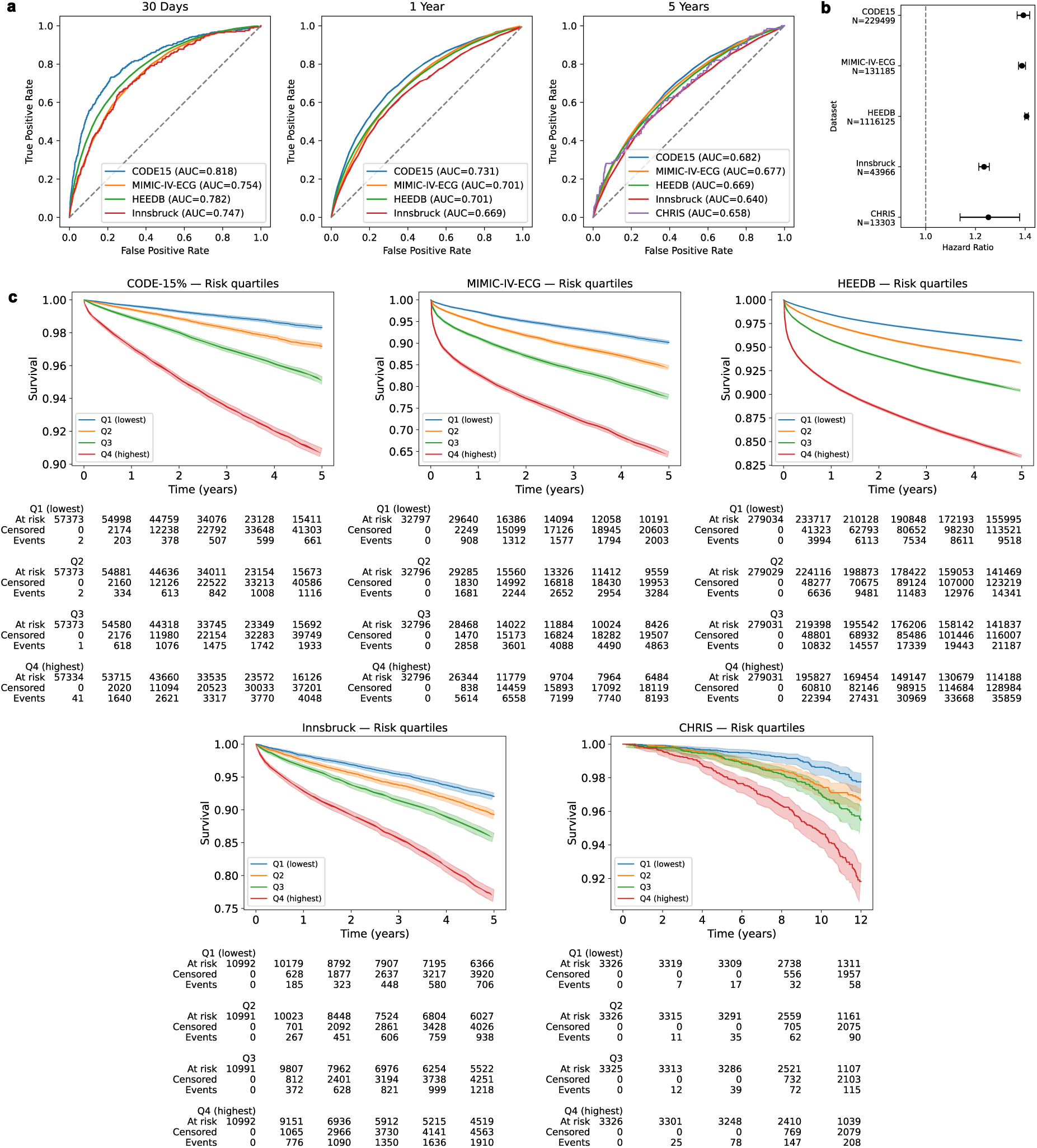
Reconstruction error is predictive of all-cause mortality. **a** Time adjusted ROC curves for 30-day, 1-year and 5-year all-cause mortality using reconstruction error. Due to a lack of events, the ROC curves for CHRIS are only shown for 5-year mortality. **b** Forest plot of hazard ratios (95% CI) for the normalised reconstruction error adjusted for age and sex for each cohort. **c** Kaplan-Meier curves for quartiles of reconstruction error for cohort. The y axis scales are different for each Kaplan Meier curve due to the wide range of event rates between cohorts. The CHRIS cohort is plot over a greater timescale due to the low event rate and long follow-up duration.

The prognostic value of the continuous reconstruction error was greatest for shorter-term outcomes (Table 2). For example, in HEEDB the age and sex adjusted hazard ratios for 30-day, 1-year, and 5-year mortality were 2.00 (95% CI 1.97-2.02, p<0.001), 1.61 (95% CI 1.60-1.62, p<0.001), and 1.46 (95% CI 1.46-1.47, p<0.001), respectively. These hazard ratios are for a change of 0.071 in reconstruction error (the standard deviation of CODE-15%). Because the hazard is multiplicative, larger differences in reconstruction error correspond to substantially larger differences in risk. For example, the 30-day mortality risk of a HEEDB participant at the 90^th^ percentile of reconstruction error (0.516) is approximately six times that of a participant at the 10^th^ percentile (0.334). These values differ by 2.6 SD, and the estimate assumes a log-linear association across this range. Figure 2a shows that time-dependent ROC analysis area under the curve (AUC) values were consistently higher for shorter prediction horizons across all cohorts, with mean AUCs of: 0.776, 0.701, and 0.667, for 30-day 1-year and 5-year mortality, respectively. Kaplan-Meier curves demonstrated clear separation between the quartiles of reconstruction error across all cohorts, with divergence across all four quartiles apparent from early in the follow-up period for all cohorts apart from CHRIS (Figure 2c).

**Table 2:** Reconstruction error is more predictive of shorter-term mortality. Age and sex adjusted hazard ratios for each cohort for 30-day, 1-year, and 5-year mortality. 95% confidence intervals for hazard ratios are shown in brackets. Each hazard ratio is the age and sex adjusted hazard for an increase of one standard deviation of reconstruction error on CODE-15%.

| Cohort | 30-day mortality |  | 1-year mortality |  | 5-year mortality |  |
| --- | --- | --- | --- | --- | --- | --- |
|  | Hazard Ratio | p | Hazard Ratio | p | Hazard Ratio | p |
| CODE-15% | 2.20 (2.07-2.34) | <0.001 | 1.60 (1.56-1.65) | <0.001 | 1.41 (1.38-1.44) | <0.001 |
| MIMIC-IV-ECG | 1.76 (1.72-1.81) | <0.001 | 1.52 (1.50-1.54) | <0.001 | 1.43 (1.41-1.45) | <0.001 |
| HEEDB | 2.00 (1.97-2.02) | <0.001 | 1.61 (1.60-1.62) | <0.001 | 1.46 (1.46-1.47) | <0.001 |
| CHRIS | — | — | 1.54 (0.86-2.75) | 0.147 | 1.36 (1.12-1.64) | 0.002 |
| Innsbruck | 1.67 (1.53-1.83) | <0.001 | 1.37 (1.32-1.43) | <0.001 | 1.26 (1.23-1.29) | <0.001 |

As a sub-analysis, participants were split into high and low risk groups using the externally derived CODE-15% threshold (normalised reconstruction error above 1.0). High-risk participants comprised 1.8% to 25.3% of each cohort (Supplementary Table S1). Elevated risk was associated with increased all-cause mortality in MIMIC-IV-ECG (HR 2.00, 95% CI 1.95-2.05, p<0.001), HEEDB (HR 1.95, 95% CI 1.93-1.98, p<0.001), Innsbruck (HR 1.53, 95% CI 1.46-1.59, p<0.001), CODE-15% (HR 1.93, 95% CI 1.84-2.02, p<0.001), CHRIS (HR 1.42, 95% CI 1.04-1.95, p=0.030), and UK Biobank (HR 1.27, 95% CI 1.08-1.50, p=0.004), the last of which was assessed using the binary threshold only. Sensitivity analyses showed hazard ratios were stable across thresholds from 0.35 to 0.55 in raw reconstruction error (Figure S5), with the chosen threshold (0.477) close to the optimum, despite not being optimised for it.

Multiple sub-analyses were performed on the MIMIC-IV-ECG dataset to explore the effect of different ECG and patient characteristics on the prognostic value of reconstruction error (Figure 3a). All sub-analyses found higher reconstruction error to be significantly associated with increased mortality. The age and sex adjusted hazard ratios for the subset of patients with and without conduction defects were 1.30 (95% CI 1.27-1.33, p<0.001) and 1.50 (95% CI 1.48-1.52, p<0.001), respectively, indicating that reconstruction error is less prognostic in patients with conduction defects. The remaining sub-analyses had similar hazards to the full cohort (1.39-1.41, compared to 1.39 in the full cohort).

**Figure 3:**
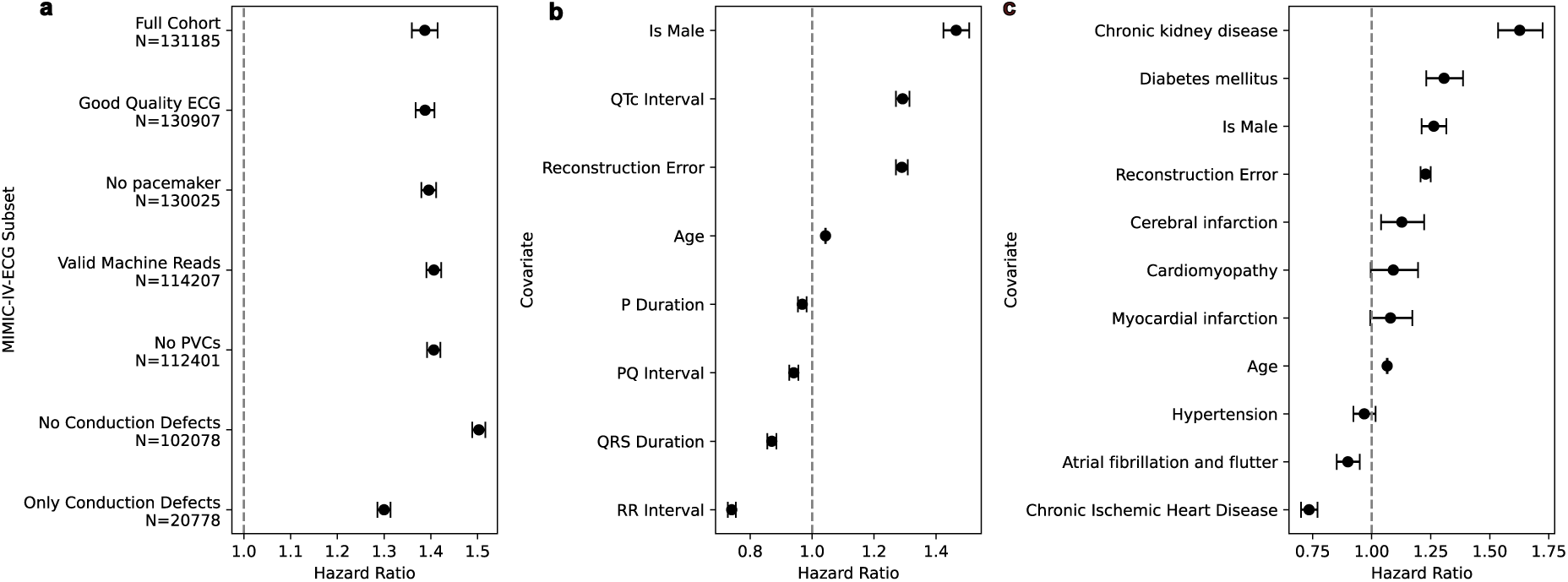
Reconstruction error remained predictive across subsets and after adjustment for additional covariates. Forest plots showing hazard ratios (95% CI) for: **a** reconstruction error adjusted for age and sex, across different subsets of MIMIC-IV-ECG; **b** a cox model adjusted for z-normalised machine-read ECG-parameters, in the subset of MIMIC-IV-ECG with valid machine reads; and **c** a cox model adjusted for comorbidities, in the Innsbruck cohort. Hazard ratios for reconstruction error and the ECG parameters represent the increase in hazard per one standard deviation increase in the respective covariate; for age, the increase in hazard per additional year.

In the MIMIC-IV-ECG cohort with valid machine-read ECG parameters, the reconstruction-based risk score remained significantly associated with mortality after adjustment for a comprehensive set of standard machine-read ECG measures (including QTc interval, QRS duration, PQ interval, P-wave duration, and RR interval), demonstrating that the reconstruction error captures prognostically relevant information beyond conventional ECG parameters (Figure 3b).

In the Innsbruck cohort, the reconstruction-based risk score remained significantly associated with all-cause mortality after adjustment for a broad panel of clinical comorbidities, including chronic kidney disease, diabetes mellitus, cardiomyopathy, cerebral infarction, myocardial infarction, hypertension, atrial fibrillation, and chronic ischaemic heart disease (Figure 3c).

Analysis across models trained for different lengths of time revealed a non-linear relationship between reconstruction quality and prognostic performance, with maximal hazard ratios observed at intermediate reconstruction quality (Table 3, Figure 4, Figure S7). Early on in training, the model has not learnt to reconstruct the ECG and so the reconstruction error is high and hazard ratios are low. However, as the model is trained for longer the reconstruction error continues to improve while the prognostic performance plateaus and then reduces (Figure 4). Table 3 shows the hazard ratios for each cohort for a model trained for 2600 steps (less than one epoch, the primary results), 15 epochs, and 100 epochs. A similar pattern was observed across models of different capacities (Figure S4), with larger models performing better at ECG reconstruction, but worse at mortality prediction. For this reason, we used the smallest model (ViT-Tiny) as it is both faster to run and provides better risk stratification.

**Figure 4:**
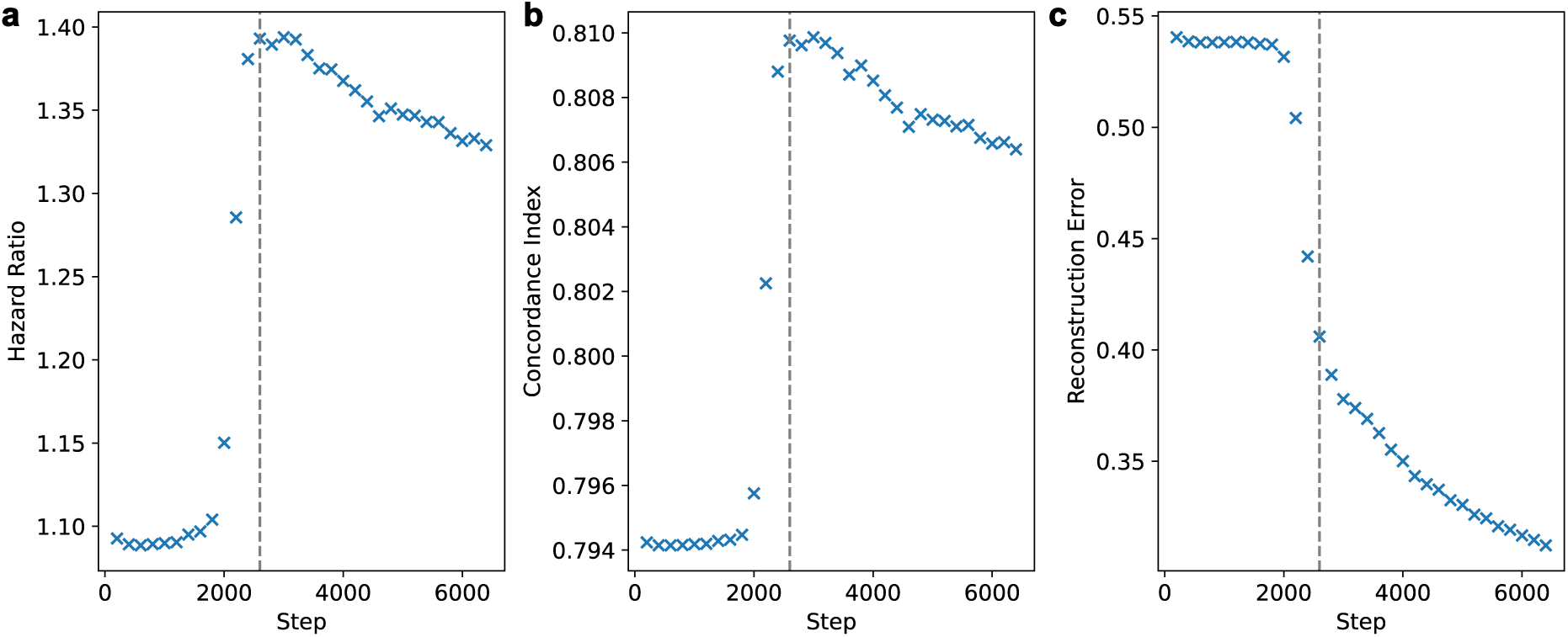
Mediocre reconstruction leads to better survival prediction. The **a** age-sex adjusted hazard ratio, **b** age-sex adjusted concordance index, **c** mean absolute error against training step for CODE-15% for the first 2 epochs of training (6400 steps). The grey line indicates the primary model (2600 steps).

**Table 3:** Hazard ratios for each cohort for models trained for different lengths. The primary model used in the paper was trained for less than 1 epoch (2600 training steps) but models trained on more data (up to 100 epochs) were evaluated for their prognostic capabilities. 95% confidence intervals for hazard ratios are shown in brackets. Each hazard ratio is the age and sex adjusted hazard for an increase of one standard deviation of reconstruction error on CODE-15%.

| Cohort | 2600 steps (primary model) |  | 15 epochs |  | 100 epochs |  |
| --- | --- | --- | --- | --- | --- | --- |
|  | Hazard Ratio | p | Hazard Ratio | p | Hazard Ratio | p |
| CODE-15% | 1.39 (1.37-1.42) | <0.001 | 1.29 (1.27-1.31) | <0.001 | 1.25 (1.23-1.27) | <0.001 |
| MIMIC-IV-ECG | 1.39 (1.37-1.40) | <0.001 | 1.37 (1.36-1.38) | <0.001 | 1.36 (1.34-1.37) | <0.001 |
| HEEDB | 1.41 (1.40-1.41) | <0.001 | 1.21 (1.21-1.22) | <0.001 | 1.15 (1.15-1.16) | <0.001 |
| CHRIS | 1.25 (1.14-1.38) | <0.001 | 1.23 (1.11-1.37) | <0.001 | 1.23 (1.11-1.36) | <0.001 |
| Innsbruck | 1.23 (1.21-1.26) | <0.001 | 1.23 (1.20-1.25) | <0.001 | 1.20 (1.17-1.22) | <0.001 |

Multiple ablation experiments were performed to demonstrate the suitability of our approach. We show that taking the mean reconstruction error across five random crops and masks leads to a stable reconstruction error, with more crops/masks providing progressively less benefit (Figure S6). Figure S6 also reveals a substantially higher variance in reconstruction error when applying a random masking strategy as opposed to the primary analysis where each timepoint is not masked in at least one lead.

Leads III and aVL had the highest mean reconstruction error across both high and low risk participants (Figure 5b). However, most leads substantially contribute to the difference between risk groups, as determined by the ratio of mean error, with leads I, II, aVR, V1, V4, V5, and V6 having ratios above 1.4. This means the high-risk participants had on average 40% greater error in these leads than the low-risk participants. A notable exception was Lead III which had a ratio of just 1.1 (Figure 5c). Visual inspection of example reconstructions revealed that the model produces near-zero predictions for Lead III across both risk groups (Figure S8).

**Figure 5:**
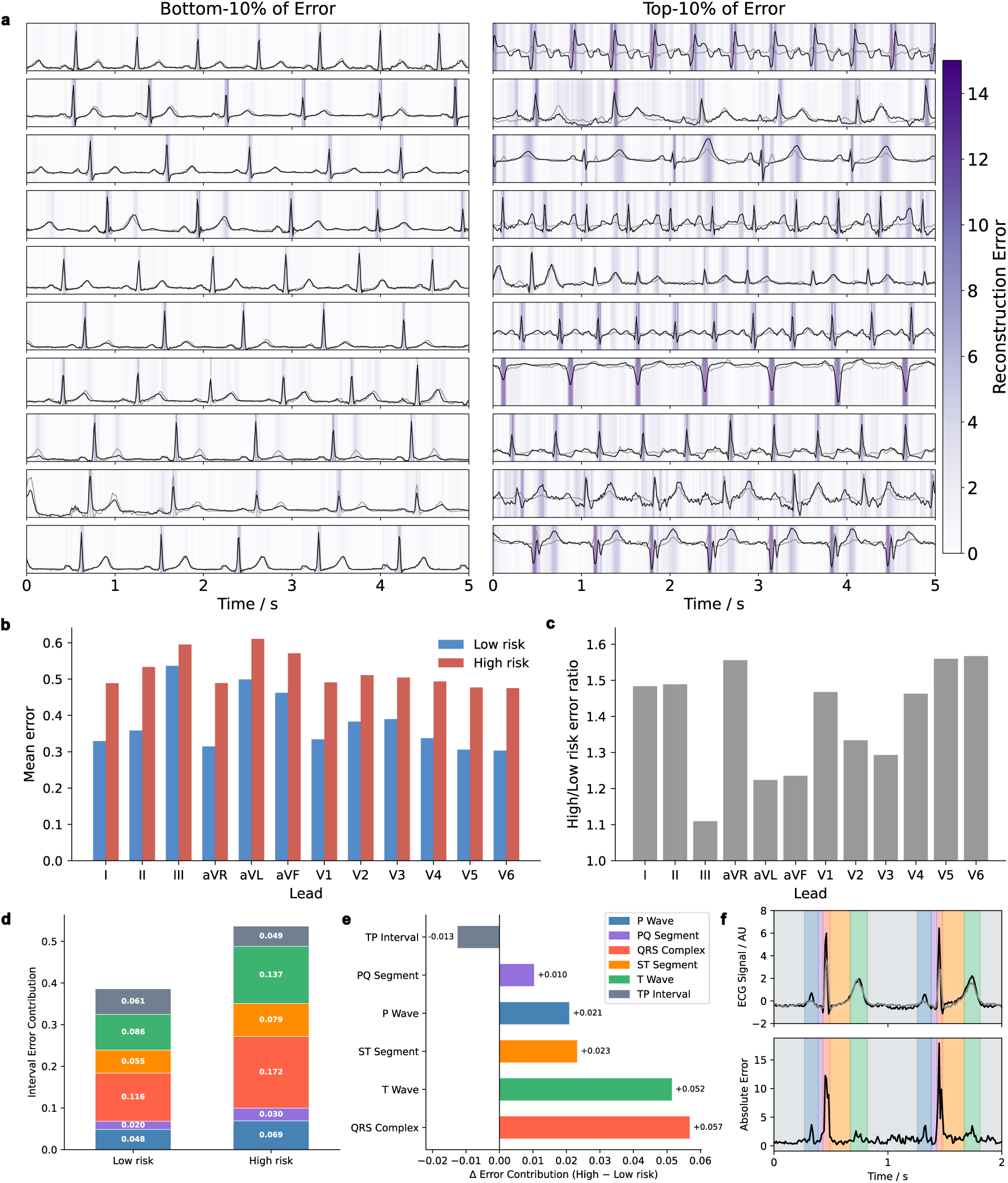
Interpretability. **a** Example Lead II ECGs (black) and reconstructions (grey) for patients in the bottom and top decile of reconstruction error in MIMIC-IV-ECG. The darker purple shaded regions highlight regions of high reconstruction error. **b** The mean contribution of each lead to the reconstruction error for low (blue) and high (red) risk patients. **c** The ratio between the high and low risk mean errors for each lead. **d** The contribution of different ECG intervals to the reconstruction error for high and low risk patients. **e** the difference in error contribution between high and low risk patients for each ECG interval. **f** An example Lead II ECG and its corresponding reconstruction and reconstruction error with interval segmentations overlayed.

All segments of the ECG contribute to the reconstruction error (Figure 5d), although the QRS complex and T wave contribute most to the difference in reconstruction error between low and high-risk participants (Figure 5e). All segments apart from the TP interval had a larger contribution to reconstruction error in high-risk participants than low risk.

Amplitude distributions were harmonised by lead-wise normalisation (Figure S2, pipelines 1 and 2). Spectral distributions were harmonised by resampling to 100 Hz (pipelines 1 and 3). At 400 Hz, cohorts differed substantially in high-frequency content, including the magnitude of powerline interference and the power retained above 40 Hz. Together with bandpass filtering to 0.5-40 Hz, these steps produced similar amplitude and spectral distributions across cohorts.

## Discussion

In this multi-cohort retrospective study, we demonstrate that a self-supervised, reconstruction-based AI-ECG risk score is a robust and continuous predictor of all-cause mortality across diverse clinical and population-based cohorts spanning critical care, hospital, and community settings on three continents. The model was trained without any labelled outcome data, relying instead on the intrinsic structure of a large clinical ECG repository. Reconstruction error from this model, reflecting the degree to which each ECG deviates from the patterns the model has learned to expect, serves as an effective and generalisable mortality risk proxy. Although model training required no labelled outcome data, the optimal training checkpoint was identified based on mortality prediction performance in a held-out validation cohort.

The prognostic value of reconstruction error was greatest for near-term mortality and attenuated over longer horizons. This is consistent with the score capturing acute deviations in cardiac electrical activity that reflect current physiological state, rather than fixed long-term baseline risk. The strong short-term signal is well suited to the high-acuity settings where early identification of deterioration is most actionable.

Optimal risk stratification was achieved not by the most accurate reconstruction, but by a model of intermediate quality, selected from less than a single epoch of training. We interpret this as evidence that the prognostically relevant signal resides in features the model finds unpredictable, i.e., features reflecting pathological deviation from normal cardiac physiology. As reconstruction quality improves, this contrast is progressively eliminated, consistent with the observation that neural networks learn simple, generalisable structure early in training before fitting more complex and dataset-specific patterns^34,35^. At this early point in training, the model has learned the broad geometry of normal cardiac electrical activity but not yet the more complex morphological patterns associated with pathology, making those patterns detectable as anomalies.

Hazard ratios were attenuated in the population-based cohorts compared with the clinical cohorts. As a continuous predictor, the association in CHRIS was weaker than in the critical care and hospital cohorts (HR 1.25 versus 1.39 to 1.41), and the same pattern held for the binary threshold in both CHRIS and the UK Biobank (HR 1.42 and 1.27, versus 1.53 to 2.00 in the clinical cohorts). This is expected, and reflects the lower absolute event rates and the healthier baseline of community-recruited volunteers. Population-level cardiovascular risk screening is precisely the setting where a scalable, low-cost ECG-based tool would be most valuable; however, this is also the setting where absolute event rates are lowest and where the clinical utility of any risk score for prevention requires prospective validation.

The reconstruction error was informative across leads covering multiple cardiac territories, suggesting it reflects generalised rather than region-specific disruption of normal cardiac electrical activity. The exception was Lead III, for which the model produces near-zero reconstructions in both risk groups, suggesting this lead had not yet been learned at the early training checkpoint selected for the primary analysis.

At the waveform level, all segments contributed to the reconstruction error, although the QRS complex and T wave had the greatest effect on the difference in reconstruction error between risk groups, indicating that both depolarisation and repolarisation abnormalities drive the risk signal. The absence of elevated error in the TP interval in high-risk patients is reassuring, suggesting the signal is carried by active cardiac electrical events rather than baseline noise or artifacts.

The robustness of the reconstruction error signal was confirmed across multiple clinically defined subgroups. PVCs generate large, morphologically irregular beats that are inherently difficult to reconstruct, and might therefore inflate reconstruction error independently of mortality risk. However, exclusion of patients with recorded PVCs produced a similar hazard ratio to the full cohort, confirming that PVC burden does not account for the prognostic signal. The conduction defect analysis is more nuanced: patients with conduction defects have higher mortality than those without, but the reconstruction error is less discriminating in this subgroup, as reflected in the substantially higher hazard ratio after their exclusion. This is likely because conduction defects (e.g., bundle branch block) alter QRS morphology in ways that inflate reconstruction error regardless of underlying mortality risk.

The preprocessing pipeline, including resampling to 100 Hz and bandpass filtering to 0.5-40 Hz, may have removed prognostic high frequency information from the ECG. However, the strong, consistent performance across five independent cohorts suggests that the retained frequency content is sufficient for mortality risk stratification. Preprocessing also served a second purpose. Reconstruction error is measured in units of signal amplitude, so between-cohort differences in amplitude would produce differences in absolute reconstruction error independent of cardiac physiology. Lead-wise normalisation removed these differences (Figure S2). Resampling to 100 Hz separately removed cohort-specific high-frequency content, including powerline interference, that reflects acquisition hardware rather than cardiac activity. Together these steps place reconstruction error on a common absolute scale, a requirement for the CODE-15% threshold to generalise across cohorts. Z-score normalisation of the reconstruction error does not achieve this, as it applies the same shift and scale to every cohort and preserves any between-cohort differences.

The reconstruction-based risk score could complement existing clinical risk assessment in several ways. As the score requires only a standard 12-lead ECG and no additional patient data, it could be computed automatically at the point of acquisition and incorporated into routine ECG reporting. In high-acuity settings, where the score showed the strongest prognostic performance, it could assist in early identification of patients at elevated short-term risk. At the population level, the score could support opportunistic risk screening, given the ubiquity of ECG acquisition in primary and secondary care. Clinical decisions are ultimately binary, and the continuous score would need to be dichotomised to trigger any specific action, such as referral or further investigation. The threshold evaluated here demonstrates that a single cut-off, derived once and applied unchanged across cohorts, identifies a high-risk group in settings ranging from critical care to population screening. The operating point appropriate for a given use would depend on the prevalence of the outcome and the relative costs of missed and unnecessary intervention, and remains to be established prospectively. Future work should assess whether the score provides prognostic information independent of, and additive to, established risk scores, and whether acting on it improves patient outcomes.

The current study has several limitations. First, the analysis is retrospective and observational; prospective evaluation is needed to establish clinical utility. Second, cause-specific mortality data was not available across all cohorts, precluding assessment of cardiovascular-specific endpoints such as sudden cardiac death or heart failure. Third, the risk score was not directly compared with established supervised AI-ECG models or conventional cardiovascular risk scores; head-to-head comparisons would be valuable to determine what complementary prognostic information the reconstruction error provides. Fourth, the binary threshold was derived in CODE-15% and applied unchanged elsewhere. Although it remained significantly associated with mortality in every cohort, the proportion classified as high risk ranged from 1.8% in CHRIS to 25.3% in MIMIC-IV-ECG. This partly reflects genuine differences in cohort mortality rates, but recalibration may be required when the target population differs substantially from the derivation cohort. Fifth, mortality follow-up in CHRIS relied on obituaries and family contact rather than registry linkage, so some deaths are likely to have been missed. A missed death is censored as a survivor. This would attenuate the observed hazard ratio, so the association in CHRIS may be stronger than reported.

The reconstruction-based framework proposed in this work is not inherently specific to the ECG and could be applied to other modalities. For example, wearable biosignals such as photoplethysmography (PPG) are a natural extension, given their continuous acquisition at scale and sensitivity to arrhythmia. More broadly, the modalities most suitable for this approach are information dense with respect to organ function. For example, our approach has been effective when applied to the ECG partly because almost all of its content reflects cardiac electrical activity, meaning reconstruction error is tightly coupled to cardiac physiology. In contrast, imaging modalities often contain large regions irrelevant to any specific physiological process, which may reduce the association between reconstruction error and health outcomes.

In summary, reconstruction error from a self-supervised masked autoencoder AI-ECG model provides a scalable, generalisable and continuous predictor of all-cause mortality. By defining risk through the inherent abnormality of the ECG signal and its deviation from the model’s physiological expectations, this approach successfully moves beyond the constraints of traditional, label-dependent algorithms. Future prospective studies will examine the clinical utility of this tool and determine its distinct prognostic value alongside established supervised AI-ECG models.

## Data Availability

The UK Biobank, CODE, MIMIC-IV-ECG, CHRIS and HEEDB datasets are available to researchers upon application to their respective data custodians. The ethics approval for the Innsbruck cohort does not allow data-sharing to external partners.

https://github.com/dlaskalab/ecg-reconstruction-risk

## Contributors

AN and CD conceptualised the study. AN, RL, SP, HB and CF accessed and verified the data. AN developed the method and AN and HB performed data analysis. RL and SP contributed to figure design. AN wrote the first draft. AB and CD critically reviewed the first draft and proposed additional experiments, which AN subsequently performed. CD supervised the project. All authors critically reviewed and commented on the manuscript. All authors had access to the data in the study and had final responsibility for the decision to submit for publication.

## Funding

This research received no specific grant from any funding agency in the public, commercial, or not-for-profit sectors and was only supported by internal funding from the Medical University of Innsbruck. The CHRIS study was funded by the Autonomous Province of Bolzano/Bozen - South Tyrol - Department of Innovation, Research, University and Museums and supported by the European Regional Development Fund (FESR1157).

## Declaration of interests

The authors declare no competing interests.

## Data sharing

Code and model weights are publicly available at https://github.com/dlaskalab/ecg-reconstruction-risk.

## Acknowledgements

This research has been conducted using the UK Biobank Resource under application number 929569. The CHRIS study thanks all study participants, the Healthcare System of the Autonomous Province of Bolzano Bozen - South Tyrol, and all Eurac Research staff involved in the study (https://www.eurac.edu/chrisack). Bioresource Impact Factor Code: BRIF6107.

## Supplementary Figures

**Figure S1:**
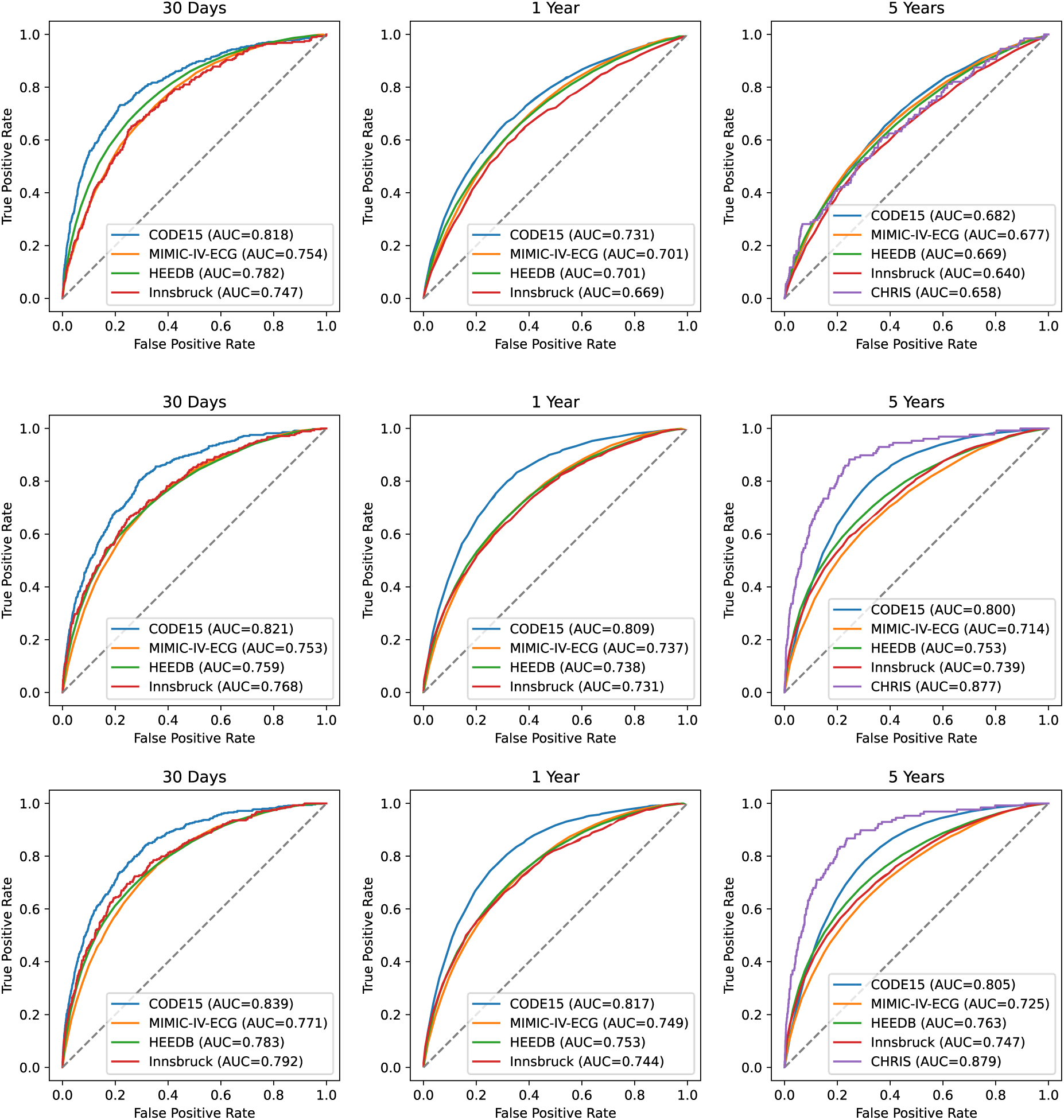
ROC curves for each cohort for 30-day, 1-year, and 5-year all-cause mortality. Row 1: Reconstruction error. Row 2: A cox model using the binarised reconstruction error, age and sex. Row 3: A cox model using reconstruction error, age, and sex. Due to low event rates, only 5-year mortality is shown for CHRIS.

**Figure S2:**
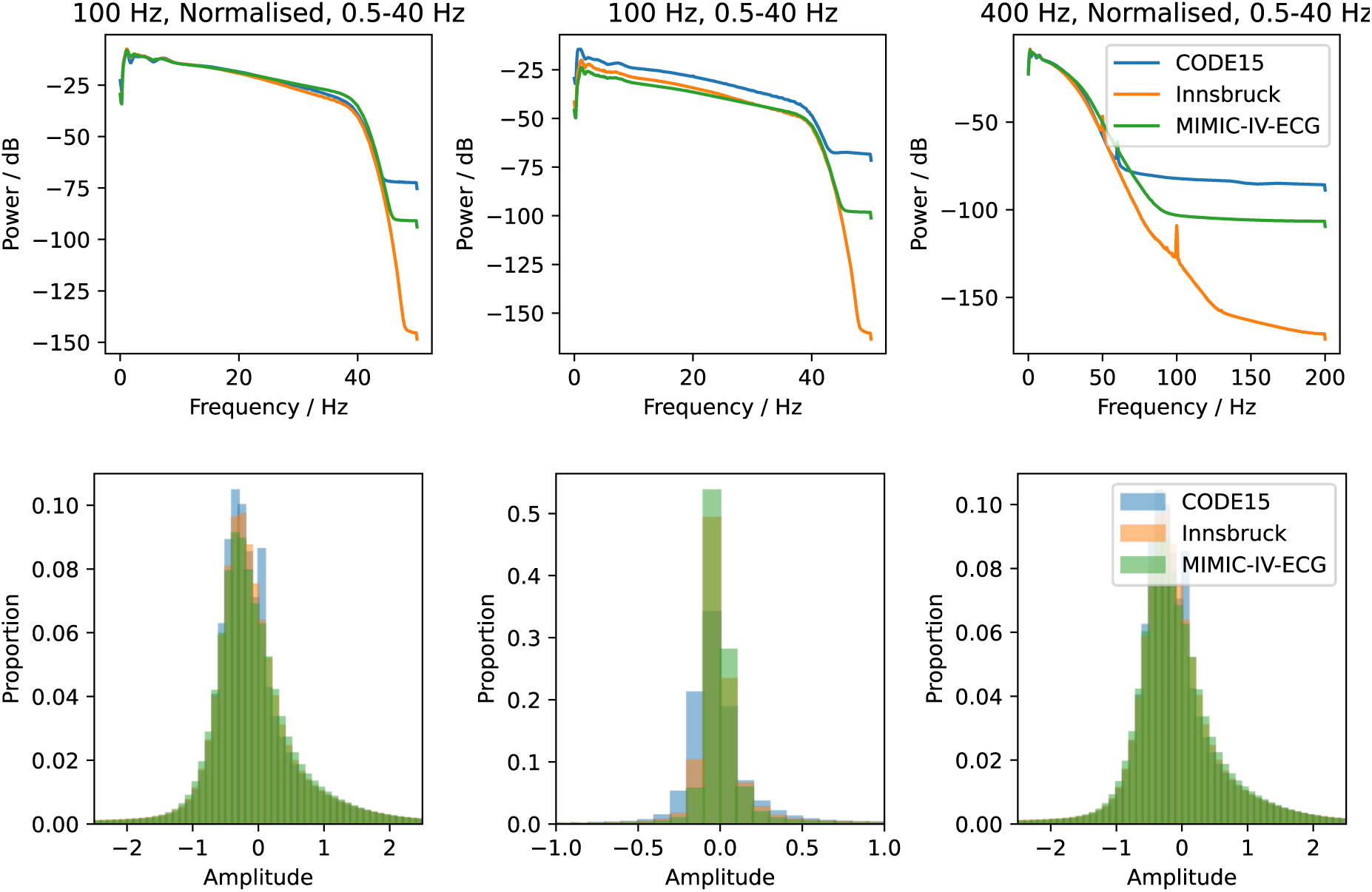
Dataset variety can be reduced with extensive preprocessing. Spectral power distributions (top) and amplitude distributions (bottom) for the CODE-15%, Innsbruck and MIMIC-IV-ECG cohorts using three different preprocessing pipelines: (1) Resampling to 100 Hz, z-normalisation and a band pass filter between 0.5-40 Hz (as in the primary analysis); (2) the same as (1) but with no normalisation; and (3) the same as (1) but resampled to 400 Hz rather than 100 Hz.

**Figure S3:**
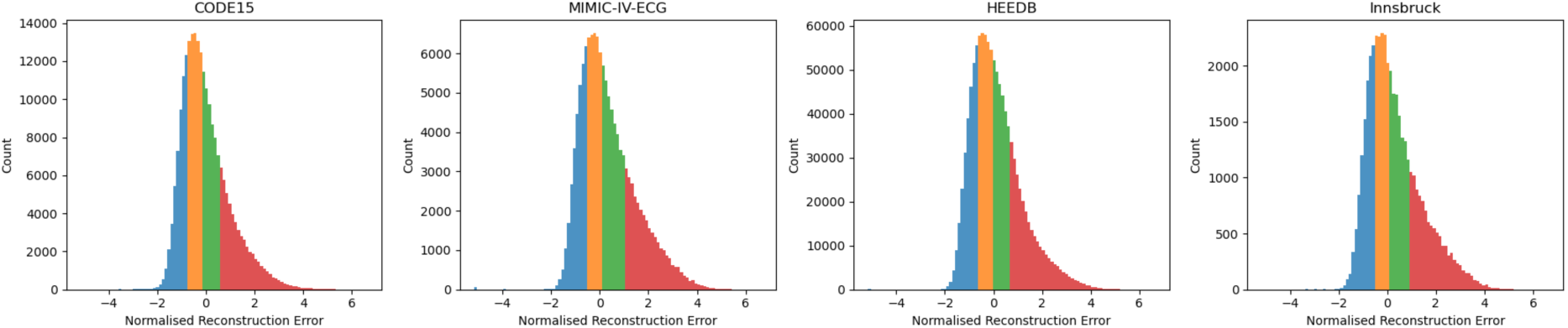
Normalised reconstruction error distributions across cohorts. Normalised reconstruction error distributions with the quartiles of error highlighted in different colours (Q1: blue, Q2: orange, Q3: green, Q4: red).

**Figure S4:**
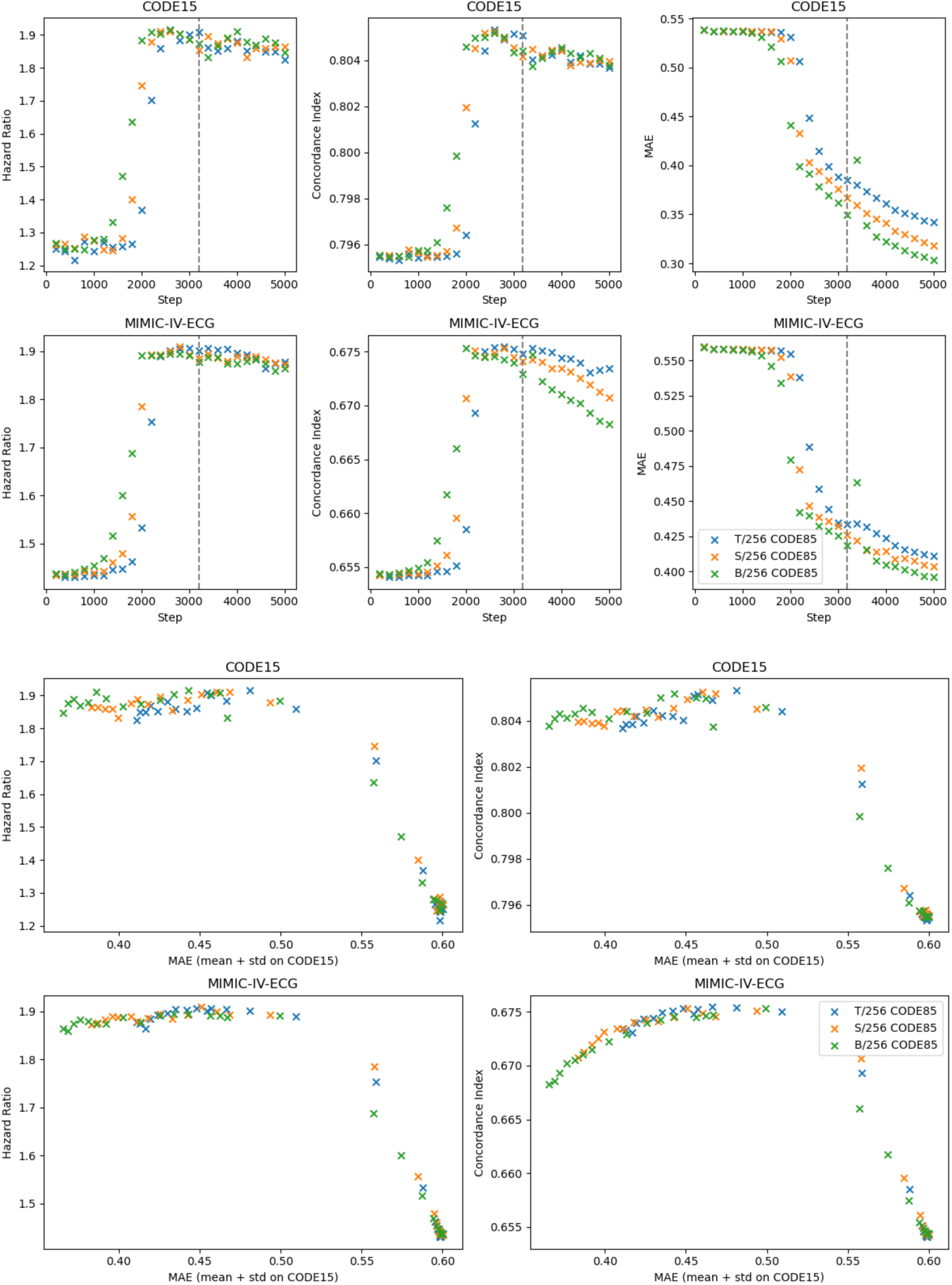
Reconstruction and mortality prediction performance for different capacity models. All models had a patch length of 256 (2.56 s) but were of varying size in terms of model parameters (ViT-Tiny, ViT-Small, and ViT-Base). The grey line indicates a single epoch of training. This analysis was performed using a binary threshold of reconstruction error of the mean + standard deviation of CODE-15%.

**Figure S5:**
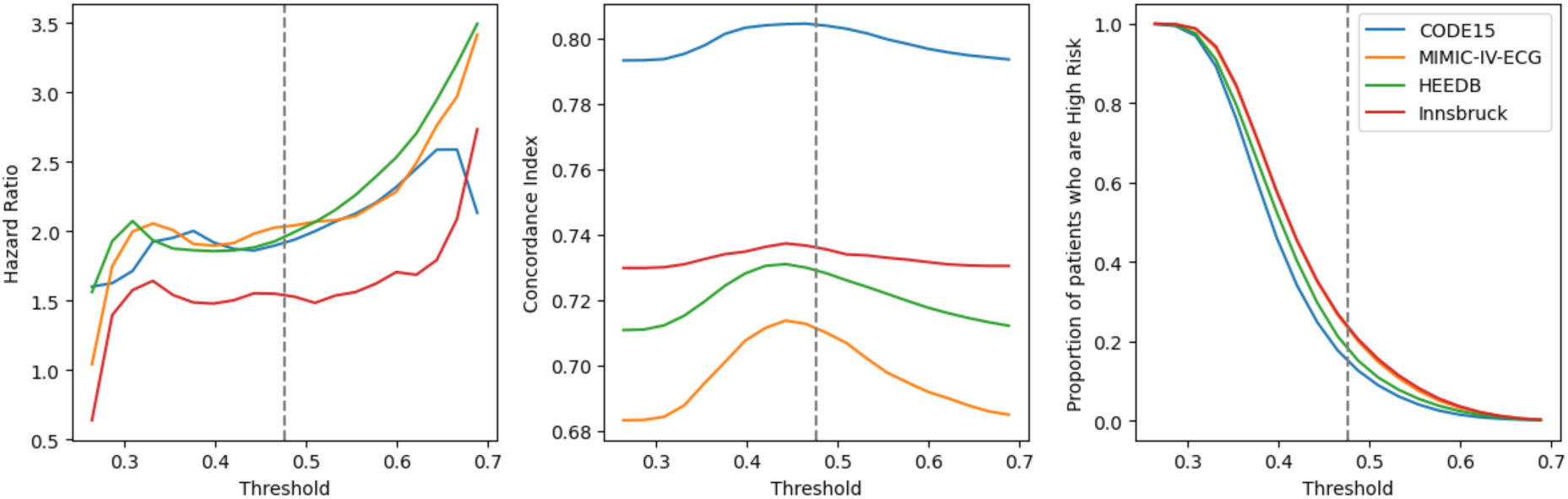
Sensitivity analysis for threshold selection. The hazard ratio (a), concordance index (b), and proportion of patients who are high risk (c), across different thresholds. The threshold used in the paper (mean + standard deviation error on CODE-15%) is visualised using a grey vertical line.

**Figure S6:**
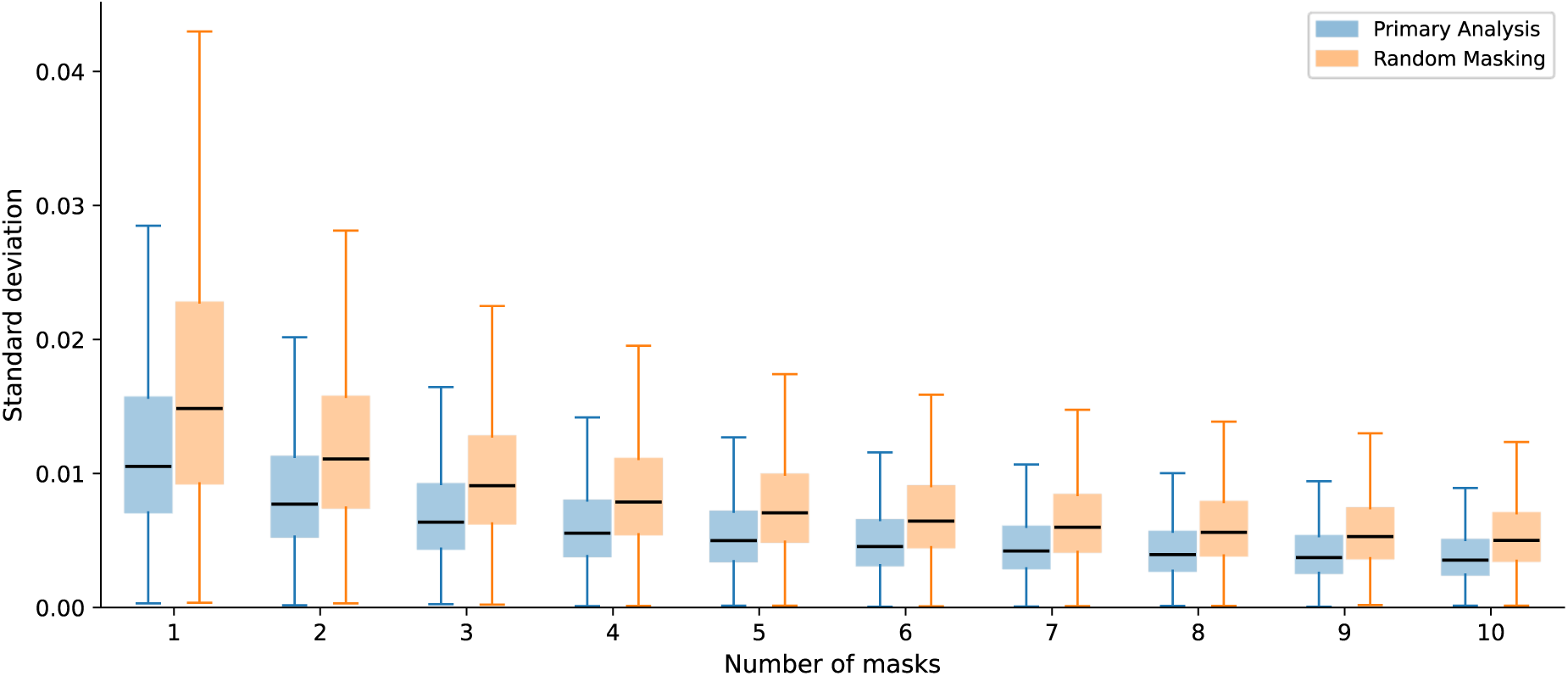
Stability of reconstruction error by number of masks averaged. Box plots show the standard deviation of mean reconstruction error across five sets of masks, for N = 1-10, computed over all recordings in the CODE-15% dataset. Each box summarises the distribution of this standard deviation across recordings. Blue boxes correspond to the primary masking strategy used throughout this paper, in which no single timepoint is fully masked across all leads, preserving spatial context at every time location. Orange boxes correspond to fully random masking, in which this constraint is lifted. Lower standard deviation indicates greater stability of the estimate

**Figure S7:**
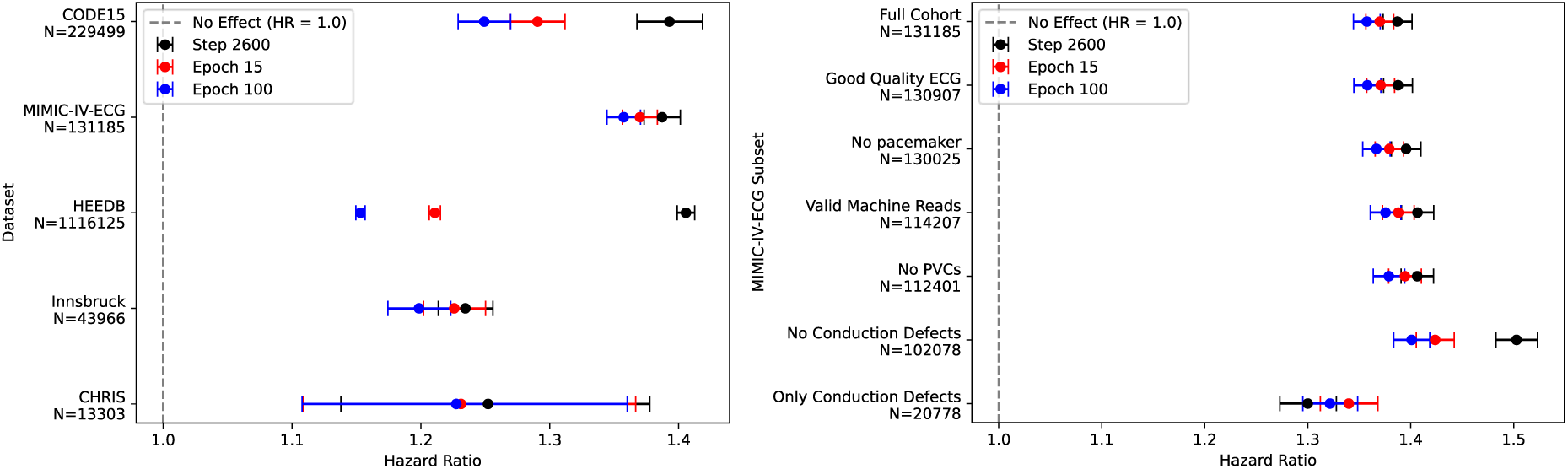
Reconstruction error across cohorts for models trained for different lengths. Forest plots showing hazard ratios (95% CI) for reconstruction error adjusted for age and sex across each cohort and for different subsets of MIMIC-IV-ECG for models trained for 2600 steps (just under 1 epoch – the model in the primary analysis – black), 15 epochs (red) and 100 epochs (blue). In each case, the hazard ratio is for an increase of one standard deviation of CODE-15% reconstruction error (0.071).

**Figure S8:**
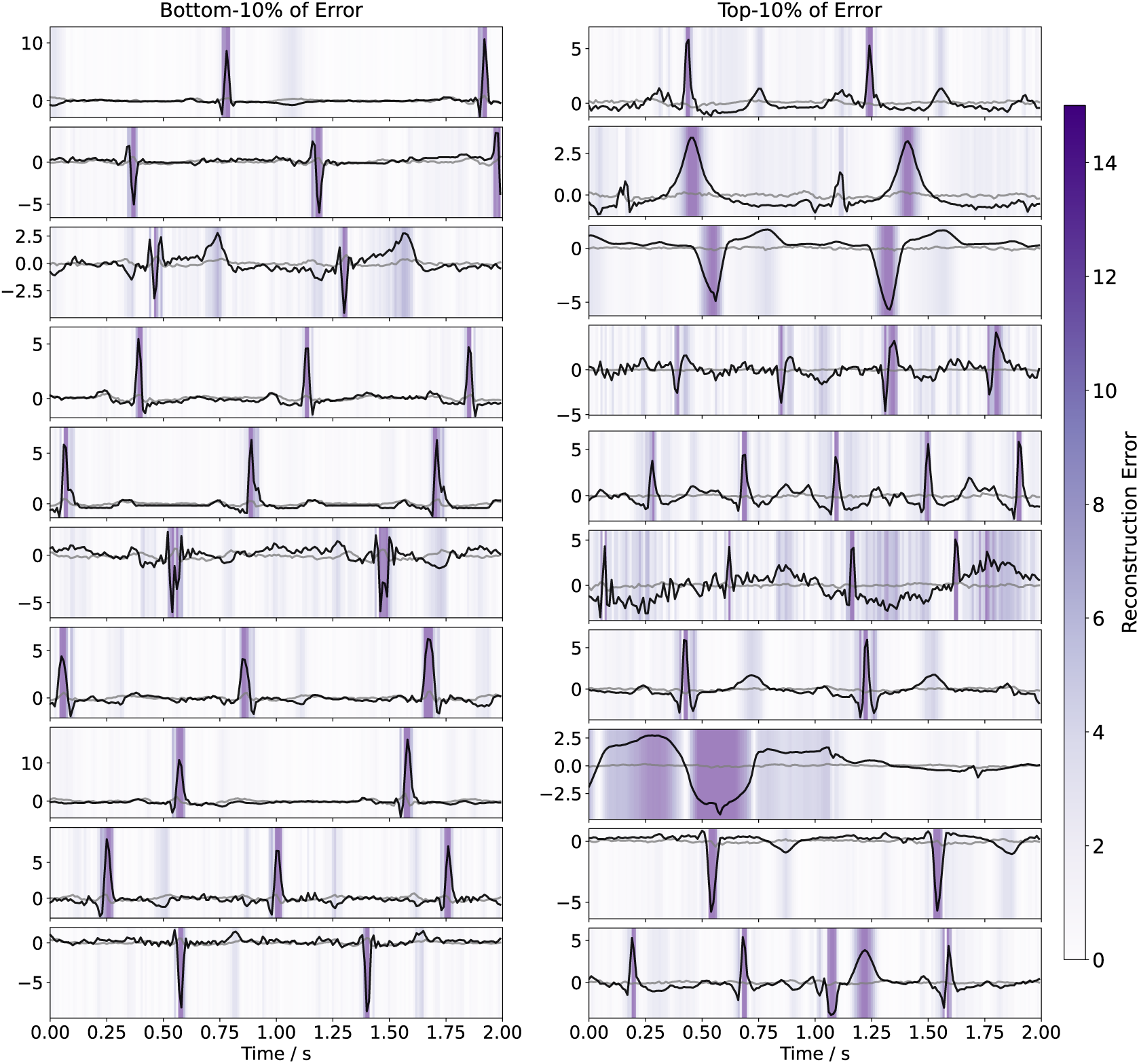
Example reconstructions for Lead III. Example Lead III ECGs (black) and reconstructions (grey) for patients in the bottom and top decile of reconstruction error. The darker purple shaded regions highlight regions of high reconstruction error.

## Supplementary Tables

**Table S1:** Demographic information for each cohort stratified by if the normalised reconstruction error is greater than 1. All values are for the cohorts after inclusion/exclusion criteria are applied. For continuous variables the median and inter-quartile range (IQR) are displayed, and statistical tests are performed using a two-sided Mann-Whitney U test. For binary variables, the number of participants and proportion of the cohort are shown, and statistical tests are performed using Pearson’s chi-squared test. Due to low numbers, and hence a risk of reidentification, 30-day and 1-year mortalities for CHRIS are not reported.

| Cohort | CODE-15% |  |  | MIMIC-IV-ECG |  |  | HEEDB |  |  | CHRIS |  |  | Innsbruck |  |  |
| --- | --- | --- | --- | --- | --- | --- | --- | --- | --- | --- | --- | --- | --- | --- | --- |
|  | Low risk | High risk | p-value | Low risk | High risk | p-value | Low risk | High risk | p-value | Low risk | High risk | p-value | Low risk | High risk | p-value |
| **N (participants)** | 194,855 | 34,644 | — | 98,042 | 33,143 | — | 914,638 | 201,487 | — | 13,063 | 240 | — | 33,578 | 10,388 | — |
| **N (%)** | 84.9 | 15.1 | — | 74.7 | 25.3 | — | 81.9 | 18.1 | — | 98.2 | 1.80 | — | 76.4 | 23.6 | — |
| **Age (years)** | 49.0<br>[33.0–<br>65.0] | 66.0<br>[48.0–<br>77.0] | <0.001 | 58.0<br>[44.0–<br>70.0] | 67.0<br>[54.0–<br>78.0] | <0.001 | 53.8<br>[41.1–<br>64.4] | 61.0<br>[47.8–<br>70.9] | <0.001 | 45.3<br>[31.0–<br>57.0] | 59.0<br>[43.0–<br>72.4] | <0.001 | 59.0<br>[47.0–<br>70.0] | 68.0<br>[56.0–<br>77.0] | <0.001 |
| **Follow-up time (years)** | 3.46<br>[2.12–<br>5.21] | 3.44<br>[2.03–<br>5.30] | <0.001 | 1.62<br>[1.01–<br>5.75] | 1.05<br>[1.00–<br>3.88] | <0.001 | 5.37<br>[1.59–<br>10.6] | 3.08<br>[0.526–<br>7.68] | <0.001 | 11.0<br>[9.14–<br>12.6] | 10.7<br>[8.72–<br>12.5] | 0.058 | 6.58<br>[2.11–<br>10.8] | 3.55<br>[1.37–<br>9.38] | <0.001 |
| **Reconstruction error** | 0.381<br>[0.350–<br>0.416] | 0.521<br>[0.495–<br>0.559] | <0.001 | 0.393<br>[0.360–<br>0.427] | 0.531<br>[0.500–<br>0.576] | <0.001 | 0.388<br>[0.354–<br>0.424] | 0.523<br>[0.496–<br>0.567] | <0.001 | 0.338<br>[0.314–<br>0.366] | 0.507<br>[0.489–<br>0.535] | <0.001 | 0.392<br>[0.360–<br>0.426] | 0.530<br>[0.500–<br>0.574] | <0.001 |
| **Normalised reconstruction error** | -0.356<br>[-0.795–<br>0.145] | 1.62<br>[1.26–<br>2.17] | <0.001 | -0.189<br>[-0.647–<br>0.301] | 1.77<br>[1.33–<br>2.41] | <0.001 | -0.261<br>[-0.735–<br>0.258] | 1.66<br>[1.27–<br>2.28] | <0.001 | -0.963<br>[-1.30–<br>-0.566] | 1.43<br>[1.18–<br>1.83] | <0.001 | -0.199<br>[-0.647–<br>0.288] | 1.76<br>[1.33–<br>2.38] | <0.001 |
| **Male sex** | 77314<br>(39.7%) | 15401<br>(44.5%) | <0.001 | 46463<br>(47.4%) | 17879<br>(53.9%) | <0.001 | 422722<br>(46.2%) | 106953<br>(53.1%) | <0.001 | 5950<br>(45.5%) | 132<br>(55.0%) | 0.004 | 17551<br>(52.3%) | 5999<br>(57.7%) | <0.001 |
| **All-cause mortality** |  |  |  |  |  |  |  |  |  |  |  |  |  |  |  |
| **30-day** | 184<br>(0.1%) | 309<br>(0.9%) | <0.001 | 1493<br>(1.5%) | 2316<br>(7.0%) | <0.001 | 6739<br>(0.7%) | 8774<br>(4.4%) | <0.001 | — | — | — | 112<br>(0.3%) | 154<br>(1.5%) | <0.001 |
| **1-year** | 1549<br>(0.8%) | 1246<br>(3.6%) | <0.001 | 5409<br>(5.5%) | 5652<br>(17.1%) | <0.001 | 25689<br>(2.8%) | 18167<br>(9.0%) | <0.001 | — | — | — | 861<br>(2.6%) | 739<br>(7.1%) | <0.001 |
| **5-year** | 4810<br>(2.5%) | 2948<br>(8.5%) | <0.001 | 10090<br>(10.3%) | 8255<br>(24.9%) | <0.001 | 52793<br>(5.8%) | 28132<br>(14.0%) | <0.001 | 117<br>(0.9%) | 11 (4.6%) | <0.001 | 2959<br>(8.8%) | 1813<br>(17.5%) | <0.001 |
| **Complete Follow-up** | 5186<br>(2.7%) | 3106<br>(9.0%) | <0.001 | 14586<br>(14.9%) | 10074<br>(30.4%) | <0.001 | 83763<br>(9.2%) | 36470<br>(18.1%) | <0.001 | 486<br>(3.7%) | 44<br>(18.3%) | <0.001 | 6397<br>(19.1%) | 3247<br>(31.3%) | <0.001 |

## Notes

### Competing Interest Statement

The authors have declared no competing interest.

### Author Declarations

The research ethics committee of the Medical University of Innsbruck gave ethical approval for this work.

